# Transarterial Therapy for Neuroendocrine Tumors: Protocol for a Systematic Review and Network Meta-analysis

**DOI:** 10.1101/2022.11.06.22281733

**Authors:** Ali Bassir, Menelaos Konstantinidis, John T. Moon, Andrew Tran, Arun Chockalingam, Katya Ferreira, Emilie Ludeman, Peiman Habibollahi, Jeff F. Geschwind, Nariman Nezami

## Abstract

**Introduction:** Neuroendocrine neoplasms are a form of tumor that develops from a variety of neuroendocrine cell types and is most commonly found in the gastrointestinal tract. As the blood from the GI tract drains into the liver, it frequently metastasizes there. Intraarterial therapies, such as bland transarterial embolisation, conventional transarterial chemoembolisation, drug-eluting beads transarterial chemoembolisation, and transarterial radioembolization with yttrium-90 are advanced methods for limiting the burden of neuroendocrine liver metastasis tumors. Currently, there is no study that compares all of these methods. Our goal is to compare the efficacy and harm of each of transarterial therapies for stage IV neuroendocrine neoplasms by conducting this systematic review and network meta-analysis.

**Methods and Analysis:** We will search electronic databases (MEDLINE, Cochrane, Embase, Scopus, ClinicalTrials.gov) from their inception to November 2022 to find all studies that compare survival and clinical finding between any two of the following intraarterial interventions: bland transarterial embolisation, conventional transarterial chemoembolisation, drug-eluting beads transarterial chemoembolisation, and transarterial radioembolization, the latter with yttrium-90. We will include randomized controlled trials (RCT), cluster-RCTs, and observational studies. We will investigate selected articles regarding our primary outcomes (Overall survival, Progression-free survival, death) and also our secondary outcomes (post embolization syndrome, hepatic abscesses, hepatic failure, gallbladder necrosis, non-target embolization). We will also search trial registries and the references of included studies to identify any additional published or unpublished studies for possible inclusion. Risk of bias will be assessed using the tools developed by Cochrane (i.e., the Cochrane RoB 2 tool for RCTs and cluster RCTs, and ROBINS-I for observational studies). We will conduct frequentists network meta-analyses, and we will use the Confidence in Network Meta-Analysis tool to assess the confidence in the evidence for the primary outcomes.

**Ethics and Dissemination:** We seek to publish our findings through peer review with a high impact, and readers will have access to the data gathered in this study. Because the underlying research is based on systematic evaluations of available data, it does not require approval by an ethical review board. The systematic review and meta-analysis’s findings will also be presented at various conferences and seminars.

**Strengths and Limitations:**

- This meta-analysis can be used to compare different types of intraarterial therapies for NELM, which can serve as a guide for choosing the best treatment option for non-surgical candidates.
- This research will lay the groundwork for future clinical trials comparing these therapy options, which can guide clinicians with a greater degree of certainty.
- Variations in specific therapy, like differences in medications used for intervention, may not be considered and may affect this comparison.

## 1. BACKGROUND

### 1.1 Description of the Condition

Neuroendocrine tumours (NETs) are considered rare tumours and account for about 0.5 percent of all cancers with an incidence of 1 or 2 per 100,000 people in the United States, of which African Americans are most affected [1]. Neuroendocrine tumors, formerly known as carcinoid tumors, arise from neuroendocrine cell compartments found throughout the body and are most commonly found in the gastrointestinal and bronchopulmonary systems [2]. Functional tumors are tumors that produce hormones in excess with resultant clinical symptoms. In addition to serotonin-producing NETs, there are insulinomas, gastrinomas, vasoactive intestinal peptide (VIP) secreting tumors, glucagonomas, and somatostatinomas.

A broad investigation of neuroendocrine epidemiology revealed an increase in the incidence of NETs[1]. It is unclear if it reflects a true increase or, rather, increased awareness and diagnostic sensitivity. The clinical presentation and biological aspects of gut cancers such as local invasion, fibrosis, and metastatic potential differ significantly depending on the anatomical position, neuroendocrine cell(s) of origin, and secretory products.

Gastrointestinal neuroendocrine tumors (NETs) often appear with advanced disease and most commonly involve the liver. While NETs are frequently indolent tumors, individuals with an advanced illness can potentially live for many years if treated [3]. At the time of diagnosis, patients with NETs present with neuroendocrine liver metastases (NELM), affecting between 46-93% of patients [4]. Due to the common development of carcinoid syndrome, the presence of NELM is not only a significant negative prognostic factor but is also linked to a lower quality of life [5].

### 1.2 Description of the Intervention

Given that both primary NET and NELM are present at the time of diagnosis for most patients, resection of the main lesion can be coupled with partial hepatectomy and/or local ablation in individuals with low to moderate NET liver metastases. The best long-term outcomes involve hepatic NELM excision with overall five-year survival (OS) approaching 60%–80% [6]. However, only about 20% to 30% of patients are resection candidates [7]. Ablative methods can be utilized either for independent therapy (percutaneous, laparoscopic, or open procedures) or as a surgical aide in patients with hepatic metastases who are not candidates for full resection [8]. Given the hypervascularity of NETLMs, like HCC, arterial embolization techniques provide efficient treatment. Patients who are not surgical candidates receive palliative therapy with image-guided intra-arterial therapies (IAT), including bland transarterial embolization (TAE), conventional transarterial chemoembolisation (cTACE), drug-eluting beads TACE (DEB-TACE) and transarterial radioembolization (TARE) with yttrium-90 (Y90) playing a vital role.

### 1.3 How the Intervention Works

Intra-arterial liver-directed treatments address NELM’s hyper-vascular characteristics. Intra-arterial treatments have been shown to be effective in the treatment of unresectable NELM [9]. They have a good safety profile as well and are now widely recommended for neuroendocrine tumors with hepatic-predominant disease [10].

As a type of IAT, the goal of bland embolization with calibrated particles is to cut off blood flow to NET metastases, depriving tumor cells of oxygen and causing cell death. A wide range of calibrated product embolization for bland TAE can be used [11]

cTACE includes transcatheter arterial delivery of one or more cytotoxic medicines combined with Lipiodol [12]. This results in a dual mechanism of tumor cell death: the chemotherapy’s cytotoxic effect combined with the embolic agent’s ischemia effect.

DEB-TACE, on the other hand, involves loading the chemotherapeutic agent onto miniature microspheres. Doxorubicin, mitomycin, cisplatin, and streptozoticin are some of the most commonly utilized drug s[13]. DEB-TACE allows for efficient delivery of the chemotherapeutic agents compared to lipiodol. [14,15].

Transarterial radioembolization (TARE) is an intra-arterial brachytherapy technique in which yttrium-90 (Y90), a beta emitter, is implanted in small microspheres and administered into the arterial blood supply of liver tumors [16]. Radioembolization can distribute large doses of radiation to hepatic NET metastases selectively, leading to a promising imaging response rate and clinical improvement.

### 1.4 Why it is important to conduct this systematic review

Currently, there are no comparisons of all the aforementioned IATs. As such, no conclusive recommendation can be made to guide clinical treatment. In a study for comparison of different types of IAT like cTACE, DEB-TACE and yttrium-90 radioembolisation, patients who had cTACE lived considerably longer than those who had DEB-TACE or Y90, although the safety and toxicity characteristics of all were equivalent [4]. In a study by Fiore et al. [17] for comparison between bland embolization and cTACE, it was shown that the median progression-free survival for all patients was 36 months in the entire population, with no notable difference between TAE and TACE. This study also demonstrated a safer profile for TAE in comparison with cTACE [17]. However, there still exists a paucity of high-quality evidence comparing the interventions in question for NELM. A multi-institutional analysis showed that there was no significant difference in median overall survival or progression-free survival between TARE and TACE patients [18]. As described, there is currently no evidence to compare all treatments for NELM to each other in the literature, and a large RCT to examine these comparisons would require considerable resources. Thus, the proposed review and network meta-analysis (NMA) will evaluate IATs for the treatment of NELM to provide guidance for stakeholders and optimize therapy for patients.

## 2. METHODS AND ANALYSIS

The present protocol has followed the guidance of the Preferred Reporting Items for Systematic Review and Meta-analysis Protocols [19] (Supplementary Material). Any changes to the protocol will be reflected in the final article and in an updated version of the PROSPERO registration. The methods section of this protocol is modeled on three previously published protocols[20-22].

### 2.1 Criteria for considering studies for this review

#### I. Type of studies

In this review, we will include any of the following study designs which compare any two interventions (see below) against one another or a control/placebo:

- Randomized or cluster-randomized controlled trials
- Observational studies

We will not exclude studies based on language, date of publication, or setting.

#### II. Type of participants

We will include studies in which adults with diagnosis of NELM (history of GI-NETs and hepatic lesions with classic radiographic features of NELM) who were not candidates for surgery and were treated using IAT.

#### III. Type of interventions

We will include studies in which any of the following interventions were compared against each other or a control:

- Bland Embolization
- cTACE
- DEB-TACE
- Y-90 radioembolization.

#### IV. Types of outcome measures

We will include studies in which any of the following outcomes are reported.

##### i. Primary outcomes

1. Overall survival (OS)
2. Progression-free survival (PFS)
3. Death

Toxicity

##### ii. Secondary outcomes

###### Minor

- Fever
- Abdominal pain
- Nausea
- Post-embolization syndrome (right-sided abdominal pain, nausea, vomiting, fever, leukocytosis, elevated liver enzymes)

###### Major

- Hepatic abscesses
- Gallbladder necrosis
- Intestinal ischemia
- Liver insufficiency

### 2.2 Search Strategy for Identification of Studies

A systematic literature search will be conducted to identify all published and unpublished trials that may be relevant to this review. The search approach created for MEDLINE via PubMed (Supplementary Material) will be adapted for searches in four different electronic database systems:

- Embase (embase.com): 1947 – Present
- Scopus (scopus.com): 1788 – Present
- Cochrane (cochrane.org): 1996 – Present
- ClinicalTrials.gov: 2008 – Present

No restriction will be applied to the publications’ language. As a part of our investigation, we will conduct a search of ClinicalTrials.gov for unpublished and ongoing trials (Supplementary Material). In addition, we will check all of the research references for any relevant articles.

### 2.3 Data Collection

#### I. Selection of studies

All references from the search strategy will be imported into a citation manager and duplicates will be removed. Subsequently, all remaining references will be uploaded into Covidence [23]. Titles and abstracts will be independently screened by two reviewers for potential inclusion. The full text of the studies included from the title-abstract screening will be obtained and two authors will independently conduct a full-text assessment with reference to the inclusion and exclusion criteria of the review (see above). We will keep records of the reason for exclusions at the full-text screening stage. Disagreements between reviewers will be resolved by discussion at any stage. If an agreement cannot be reached, a third reviewer will make the ultimate decision on inclusion as necessary. The results of the critical appraisal will be displayed in a PRISMA flowchart [24].

#### II. Data extraction and management

Using a standardized extraction form, two reviewers will independently extract data from each of the included studies. Any disputes will be addressed by discussion or, if required, by a third reviewer, as in the critical evaluation. One author will enter relevant extracted data into Review Manager 5 [25] and a second author will double-check it for correctness. After that, the data will be exported to R [26]. We will extract information on study characteristics, outcomes, and possible impact modifiers from each study, to the extent that it is accessible. We will reach out to the corresponding author of a study if any information is missing or if further information is required (e.g., for risk of bias assessment).

##### i. Study characteristics data

The following information is extracted from articles: The article title, corresponding author’s name, type of publication, publication date, research setting, conflicts of interest, study design, funding sources, patient features (e.g., age and gender), study-level inclusion/exclusion criteria, and how missing data was dealt with in the analysis (if applicable).

##### ii. Outcome data

Compared interventions, as well as all primary and secondary results measures, will be collected along with the corresponding measure of uncertainty. For outcomes and baseline characteristics, we will try to collect arm-level data from RCTs or study-level data if this is not possible. For observational studies, we will collect adjusted effect estimates or, if they aren’t available, unadjusted effect estimates, together with a measure of uncertainty for each.

##### iii. Potential Effect modifier data

We will also extract the rates of attrition, trial size, tumor type, ECOG status, liver replacement percentage, and hepatic tumor burden.

### 2.4 Risk of bias

Each study included in this systematic review will undergo a risk of bias assessment according to Cochrane Handbook for Systematic Reviews of Interventions [27]. Two reviewers will assess the risk of bias independently using the Cochrane’ Risk of Bias’ tool for randomized trials (RoB 2) [28,29] and non-randomized studies using Risk Of Bias In Non-randomised Studies of Interventions (ROBINS-I) [30]. If the reviewers are unable to reach an agreement, a third reviewer will address any potential conflicts.

For RCTs, the risk of bias will be categorized as either ‘low’, high,’ or ‘some concerns,’ and will be assessed for each of the following five domains:

1. Bias resulting from the randomization process;
2. Bias resulting from deviations from the intended interventions;
3. Bias resulting from missing outcome data;
4. Bias in outcome assessment; and
5. Bias resulting from the selection of the reported findings

For non-randomized studies, the risk of bias will be categorized as either Similarly, using the ROBINS-I tool, we will assess the risk of bias in non-randomized studies for against each of the following domains:

1. Bias owing to confounding
2. Bias in the study’s participant selection
3. Bias in intervention classification
4. Bias as a result of interventions that deviate from the planned interventions
5. Bias as a result of missing data
6. Bias in the outcome measurement
7. Selection bias in the stated result

For risk of bias assessments conducted using either the RoB 2 or ROBINS-I tool, the risk of bias for each domain and the overall risk of bias will be assessed using a set of signaling questions that are part of the tools.

### 2.5 Data synthesis

#### I. Characteristics of included studies

We will summarize all of the included studies through descriptive statistics. Additionally, we will report on the population characteristics for each study, including the comparison arms and study design. For each outcome, a network diagram will be presented, providing a graphical representation of the body of evidence for a given outcome with edges proportional to the number of studies per comparison and nodes proportional to the number of patients (Figure 1). An edge is drawn between nodes only if a study is included in which the comparison is made between interventions corresponding to the nodes in question.

**Figure 1.**
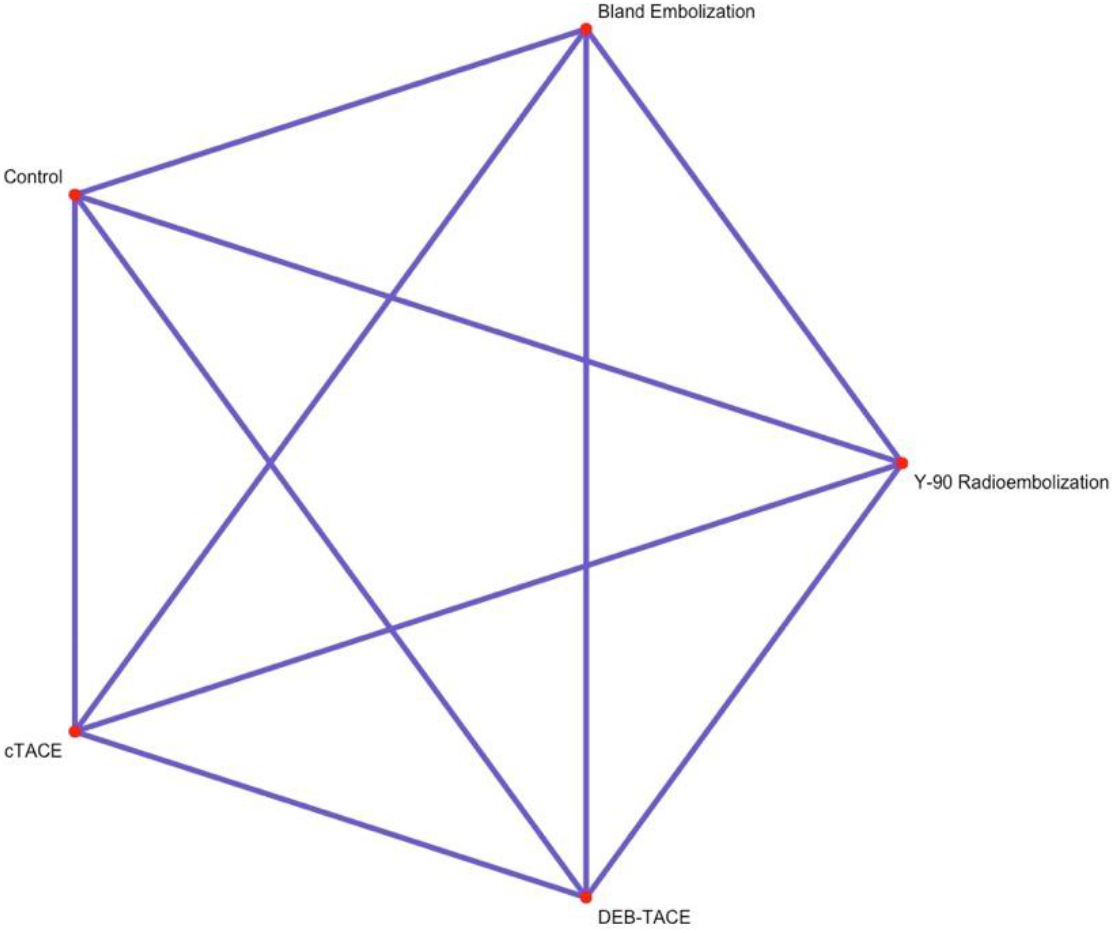
Example of a network diagram with interventions considered in this review.

#### II. Relative treatment effect

We will estimate pairwise relative treatment effects through the relative risk (RR) for dichotomous outcomes, hazard ratios (HR) for time-to-event or survival outcomes, and mean difference (MD) or standardized mean difference (SMD) for continuous outcomes. For each outcome, we will also provide a ranking among the set of included interventions, estimated using the p score – a scalar value from 0 (denoting the least effective intervention) to 1 (denoting the most effective intervention) [31].

#### III. Pairwise meta-analysis

Given more than one study, a pairwise meta-analysis will be conducted for each pair of comparisons using an inverse-variance weighted random effects model. We will conduct the estimation using the restricted maximum likelihood estimator. To account for the possibility of small trials, we will estimate the summary effect estimates using the Hartung-Knapp-Sidik-Jonkman method [32,,33]. Analyses will be carried out in R [26] using the meta package[34].

#### IV. Network meta-analysis

Due to the possible methodological and clinical heterogeneity of potentially included studies, we will fit random effects network meta-analysis models for each outcome. In fitting the models, we will account for the correlation from multi-arm trials. We will assume a common between-study variance. We anticipate that both RCTs and non-randomized studies (NRS) will be included; as such, for each outcome, we will:

1. Fit an NMA only the included RCTs
2. Fit an NMA only to the included NRS
3. Fit an NMA to all included studies (RCT and NRS)
4. Fit a “design adjusted” NMA and hierarchical NMA [35]

#### V. Assessment of heterogeneity, transitivity, and statistical inconsistency

##### i. Clinical and methodological heterogeneity

For pairwise meta-analyses, heterogeneity will be estimated by the I^2^ statistic and by visual inspection of forest plots [36,37]. Furthermore, we will consider any meaningful clinical groupings that may account for heterogeneity.

##### ii. Statistical Inconsistency

We will use the “back-calculation method” to assess local inconsistency [38]. Furthermore, to assess global inconsistency, we will use the “design-by-treatment interaction” model [39]. In networks with RCT and NRS studies, we will assess differences between the four different types of evidence (i.e., direct randomized, direct non-randomized, indirect randomized, and indirect non-randomized) [35]. Discrepancies between the four types of evidence will be further investigated, and if a source of disagreement is identified, we will try to explore it through meta-regression or subgroup analysis [40].

##### iii. Transitivity

Lastly, to assess the transitivity assumption (a key assumption in NMA) is that the distributions of effect modifiers do not differ across treatment comparisons. In this regard, to assess possible violation of the assumption, we will compare the distribution of potential effect modifiers across each pairwise comparison using direct evidence (where possible). We will report on any meaningful violations of the transitivity assumption.

### 2.6 Unit of analysis issues and missing data

We will include multiarm trials in the review. Additionally, we will account for the correlation between effect estimates in the networks. Cluster and cross-over RCTs are not expected; however, if any such studies are included, we will proceed as recommended in the Cochrane Handbook for Systematic Reviews of Interventions [27]. We will carry out all analyses on an intention-to-treat basis.

Missing data for each included study will be investigated (e.g., the extent of the missing data and whether this was adequately addressed). A sensitivity analysis will be conducted for primary outcomes such that studies with substantial missing data are excluded. The severity of the degree of missing data will be evaluated on a case-by-case basis, and a threshold for “substantial missing data” will be established based on the set of included studies. Lastly, where possible, measures of uncertainty will be reported in terms of standard errors, and if uncertainty cannot be ascertained, we will impute using the mean of the standard errors from the included studies [41,42].

#### I. Assessment of reporting biases

For the primary outcomes, provided that we have at least 10 included studies, we will visually inspect the funnel plots for each treatment. Moreover, we will assess comparison-adjusted funnel plots to investigate whether there is any evidence of publication bias [40]. Where possible, we will conduct a meta-regression with the study variance as the covariate [43].

### 2.7 Exploring heterogeneity and inconsistency

For the primary outcomes, we will investigate possible sources of heterogeneity (i.e., ECOG status, liver replacement percentage, hepatic tumor burden, and tumor type). Given sufficient studies, subgroup analyses will be conducted based on the above sources of heterogeneity. Furthermore, should sufficient studies be available, we will conduct a sensitivity analysis based on:

Missing data (i.e., exclusion of studies will result in a substantial degree of missing data)

Study size (we will restrict small studies as they may lead to publication bias). Small studies will be defined as those that are smaller than the median study size of the included studies.

### 2.8 Assessment of confidence in network estimates and summary of findings

We will create a ‘summary of findings’ table to present the evidence comparing treatments. Outcomes in the table will include:

1. Overall survival
2. Progression-free survival
3. Death
4. Toxicity

We will assess the confidence in the findings using the Confidence in Network Meta-Analysis (CINeMA) frameworks [44,45] across the following domains to arrive at a final rating of the confidence of the evidence:

1. Within-study bias
2. Across-study bias
3. Indirectness
4. Imprecision
5. Heterogeneity
6. Incoherence

## 3. CONCLUSIONS

### 3.1 Limitations

There are restrictions on the scope of the proposed research. Bland Embolization, cTACE, DEB-TACE, and Y-90 TARE are the only categories we can use as we are using aggregated data for our planned meta-analyses rather than individual patient meta-analysis. This implies that we likely not be able to take into account differences in a specific therapy within and between studies. As a consequence of comparing numerous treatments, we may discover statistically significant findings simply by coincidence; yet, there are no well-established techniques in systematic reviews to account for this problem. With this in mind, we think that although the findings are useful, interpreting them should be done carefully.

### 3.2 Summary

Although NELM tumors are rare, their mortality rate is high. Treatments that have higher survival rates and lower complications can significantly benefit these patients. This systematic review and meta-analysis aimed at comparing different types of intra-arterial therapies including Bland Embolization, cTACE, DEB-TACE, and Y-90 TARE for the treatment of neuroendocrine tumor liver metastasis. Understanding the efficacy and safety of these therapies can lead to enhanced judgment of physicians in tailoring therapy for these patients. Therapies with lower complications can benefit patients with comorbidities, as management of complications in these cases would be challenging. In addition, the cost-benefit of treatments can be considered in treatment.

### 3.3 Ethics and dissemination

This research did not need to be approved by an ethical review board because it is based on systematic analyses of existing data. The results of the systematic review and meta-analysis will be presented at conferences and seminars, as well as published in a respectable peer-reviewed journal.

## Data Availability

All data produced in the present work are contained in the manuscript.

## 3.6 Funding

There was no specific grant for this research from any funding body in the public, private, or not-for-profit sectors.

### 3.7 Patient and public involvement

There were no patients or members of the public who were engaged in the development of this protocol, and they will not be involved in the finalization or dissemination of the review and results.

